# Incidence of pancreatic cancer in angiotensin-converting enzyme inhibitors (ACEIs) versus angiotensin receptor blockers (ARBs): a population-based cohort study

**DOI:** 10.1101/2022.07.26.22278092

**Authors:** Gary Tse, Jiandong Zhou, Sharen Lee, Joshua Kai Fung Hung, Keith Sai Kit Leung, Ying Liu, Yuhui Zhang, Tong Liu, Wing Tak Wong, Ian Chi Kei Wong, Qingpeng Zhang, Bernard Man Yung Cheung

## Abstract

**Background:** Angiotensin-converting enzyme inhibitors (ACEIs) and angiotensin receptor blockers (ARBs) have been associated with lower rates of pancreatic cancer. However, some studies did not similarly reveal significant associations. The objective of this study was to examine the associations between use of ACEIs or ARBs and incident pancreatic cancer.

**Methods:** Patients who were prescribed ACEI or ARB between 1 January 2000 and 31 August 2020 at Hong Kong public hospitals, or their associated clinics and ambulatory care facilities were included. The primary outcome was incident pancreatic cancer.

**Results:** A total of 411,883 patients (ACEI users: n=355771, 86.38%; ARB users: 56112, 13.62%) were included. Over a median follow up of 2875 days (SD: 1901), 1194 incident cases of pancreatic cancer (0.28%) were identified. After 1:1 propensity score matching, univariable Cox regression found that ARB use was associated with lower risks of new onset new onset pancreatic cancer (hazard ratio [HR]: 0.69, 95% CI: [0.53, 0.90], P=0.0065). This association remained significant after multivariable adjustment (HR: 0.67, 95% CI: [0.52, 0.88], P=0.0036). Similar conclusions were reached on competing risk analyses using cause-specific and subdistribution hazard models before and after matching, and after inverse probability of treatment weighting. Subgroud analyses identified higher protective effects of ARB exposures in females than in males, while more severe adverse risks of ACEI exposure effects for males than for females (log-rank test, P<0.05).

**Conclusions:** ARB use was associated with lower risks of new onset pancreatic cancer both before and after propensity score matching compared to ACEI use.

## Introduction

The renin–angiotensin–aldosterone system (RAAS) is resposnible for blood pressure regulation. Medications that inhibit this system are known to exert anti-hypertensive effects, with additional benefits on reverse cardiac remodelling. Moreover, the identification of local RAAS in different tissue types has led to insights into its regulation of cell migration, proliferation and other functions. Angiotensin II also mediates survival of malignant cells in some cancer types ^1^. These findings therefore suggest a possible anti-carcinogenic effects of RAAS inhibitors on cancer.

Indeed, clinical and epidemiological studies have demonstrated better survival in cancer patients who are prescribed RAAS inhibitors ^2^. For example, a meta-analysis found that use of ACEIs or ARBs conferred benefits on all-cause, cancer-specific and recurrence-free survival for digestive malignancies ^3^. Moreover, the combination use of ACEI or ARB with the anti-neoplastic agent gemcitabine led to better survival in advanced pancreatic cancer patients ^4^. Indeed, RAAS inhibitors are increasingly explored as additional therapy for pancreatic cancer ^5^.

However, there are few studies that have investigated the relationship between long-term exposure of ACEIs or ARBs and pancreatic cancer, unlike for lung ^6^or colorectal cancer ^7, 8^. Firstly, the prescription of anti-hypertensive agents, including RAAS inhbitors, was not associated with altered risks of pancreatic cancer development in patients with established chronic pancreatitis ^9^. Secondly, lower risks of pancreatic cancer in ACEI/ARB users were found compared to patients who used calcium channel blockers ^10^. Thirdly, ARB use was found to be associated with better survival in patients undergoing surgical resection of pancreatic cancer ^11^. Finally, in an observational study using data from the United Kingdom Clinical Practice Research Datalink, ARBs or ACEIs was not significantly associated with reduced risks of pancreatic cancer compared to other antihypertensive drugs, although there was a short-term benefit of ARB use for 1-3 years on pancreatic cancer risk, but this was lost after 3 years ^12^. Given the paucity of large-scale epidemiological studies on comparative effects of pancreatic cancer outcomes in users of RAAS inhibitors, we examined the relationship between ACEI and ARB use and pancreatic cancer risk using a population-based database in Hong Kong.

## Methods

### Study design and population

This population-based retrospective cohort study was designed to compare the long-term effects of ACEI or ARB exposure on new onset pancreatic cancer risk. This study was approved by The Institutional Review Board of the University of Hong Kong/Hospital Authority Hong Kong West Cluster and The Joint Chinese University of Hong Kong – New Territories East Cluster Clinical Research Ethics Committee. The requirement for informed consent was wavied owing to the retrospective and observational nature of this study. Patients and the public were not engaged in this study. The patients were identified using the Clinical Data Analysis and Reporting System (CDARS), a city-wide database of patient information who attend public hospitals in the Hong Kong city of China. This system includes demographics such as age and gender, diagnoses made by healthcare professionals, and prescription of all medications in accident and emergency, inpatient and outpatient settings. Local teams from China have made use of CDARS to conduct observational cohort studies on comparative drug action ^13, 14^ including anti-hypertensives ^15, 16^, development of predictive models ^17^ and specific diseases ^18, 19^.

The inclusion criteria were patients receiving either ACEI or ARB prescriptions in public healthcare facilities under the management of the local hospital authority. The exclusion criteria were patients receiving both ACEI and ARB, and those with diagnoses of colorectal cancer before initial ACEI/ARB index use. The following comorbidities were extracted based on ICD-9 coding (**Supplementary Table 1**): diabetes without chronic complication, diabetes with chronic complication, hypertension, heart failure, atrial fibrillation, renal diseases, liver diseases, ventricular tachycardia/fibrillation, dementia, ischemic heart disease (IHD), acute myocardial infarction (AMI), peripheral vascular disease (PVD), stroke/transient ischemic attack (TIA), chronic obstructive pulmonary disease (COPD), gastrointestinal bleeding, obesity and cancer before initial prescription of ACEI/ARB agents.

### Outcomes and statistics

The primary outcome was new onset pancreatic cancer,. Patients were followed up from initial ACEI/ARB drug prescription (baseline date) to date of event occurrence, death or study end of follow-up until August 31st, 2020. Descriptive statistics were used to analyze the basleine characteristics of the included cohorts. Continuous variables were expressed as mean values along with their 95% confidence interval [CI] or standard deviation [SD]. Categorical variables were expressed as frequency (percentage). Comparisons for continuous variables were made using the Mann-Whitney U test. The χ^2^ test with Yates’ correction was used for 2×2 contingency data. Propensity score matching with a 1:1 ratio was used to generate a cohort of ARB and ACEI users based on baseline demographics, Charlson comorbidity index, CHA-DS-VASc score, past comorbidities, and non-ACEI/ARB medications with nearest neighbour search strategy.

Cox regression was applied to evaluate the significant risk factors and the relationship between ACEI/ARB monotherapy and incident pancreatic cancer. Competing risk analyses were conducted using cause-specific and subdistribution hazard models. Multiple approaches using the propsensity score were employed, including propensity score stratification ^20^, high dimensional propensity score adjustment ^21^ and inverse probability of treatment weighting ^22^. Hazard ratios (HRs) with their 95% CIs/P-values were calculated. P-values below 0.05 were considered statistically significant. All statistical analyses were conducted using RStudio (Version: 1.1.456), Python (Version: 3.6) or Stata (Version: SE 16.0).

## Results

### Basic characteristics

Initially, 500399 patients who received ACEI or ARB therapy were found (**Figure 1**). Patients prescribed ACEI and ARB at baseline or on follow-up (n=85563), or had any type of cancers at or before baseline were excluded. Thus, this study included 411282 patients. 56112 patients (13.62%) received ARBs and 355771 (86.38%) received ACEIs. Over 2875 days of median follow-up (standard deviation [SD]: 1901 days), 1194 new onset pancreatic cancer events (0.28%) were identified. The results of 1:1 propensity score matching between ARB (n=56112) and ACEI (n=56112) users are shown in **Table 1**. The comparisons of clinical and baseline characteristics of caregorised by incident pancreatic cancer outcomes are presented in **Supplementary Table 2**. Before matching, the ARB group had a higher CHA_2_DS_2_-VASc score (mean±SD: 2.4±1.6 vs. 1.9±1.5; standardized mean difference [SMD]=0.28) and a higher frequency of hypertension (34.85% v.s. 17.17%, SMD=0.41) and ischaemic heart disease (15.96% v.s. 7.92%, SMD=0.25). These imbalances between the two groups were reduced (SMD<0.2) after propensity score matching.

**Table 1.** Baseline characteristics of patients with ARB v.s. ACEI use before and after 1:1 propensity score matching. * for SMD≥0.20; # indicates the difference between patients with/without new onset pancreatic cancer

| Characteristics | Before matching |  |  | After matching |  |  |
| --- | --- | --- | --- | --- | --- | --- |
|  | ARB (N=56112)<br>Mean(SD);Max;N or<br>Count(%) | ACEI (N=355771)<br>Mean(SD);Max;N or<br>Count(%) | SMD | ARB (N=56112)<br>Mean(SD);Max;N or<br>Count(%) | ACEI (N=56112)<br>Mean(SD);Max;N or<br>Count(%) | SMD |
| ***Demographics*** |  |  |  |  |  |  |
| Male gender | 25753(45.89%) | 192707(54.16%) | 0.17 | 25753(45.89%) | 25808(45.99%) | <0.01 |
| Female gender | 30359(54.10%) | 163064(45.83%) | 0.17 | 30359(54.10%) | 30304(54.00%) | <0.01 |
| Baseline age, years | 69.5(13.6);110.2;n=56112 | 69.1(13.8);116.3;n=355771 | 0.02 | 69.5(13.6);110.2;n=56112 | 69.5(13.6);106.8;n=56112 | <0.01 |
| <40 | 1255(2.23%) | 9764(2.74%) | 0.03 | 1255(2.23%) | 1255(2.23%) | <0.01 |
| [40,49] | 2993(5.33%) | 21691(6.09%) | 0.03 | 2993(5.33%) | 2989(5.32%) | <0.01 |
| [50,59] | 7935(14.14%) | 47058(13.22%) | 0.03 | 7935(14.14%) | 7908(14.09%) | <0.01 |
| [60,69] | 12032(21.44%) | 69837(19.62%) | 0.04 | 12032(21.44%) | 12005(21.39%) | <0.01 |
| [70,79] | 14707(26.21%) | 105667(29.70%) | 0.08 | 14707(26.21%) | 14762(26.30%) | <0.01 |
| >80 | 14087(25.10%) | 82076(23.06%) | 0.05 | 14087(25.10%) | 14129(25.17%) | <0.01 |
| ***Past comorbidities*** |  |  |  |  |  |  |
| Charlson score | 3.1(1.8);14.0;n=56112 | 2.8(1.6);15.0;n=355771 | 0.18 | 3.1(1.8);14.0;n=56112 | 3.1(1.8);14.0;n=56112 | <0.01 |
| CHA-DS-VASc score | 2.4(1.6);9.0;n=56112 | 1.9(1.5);9.0;n=355771 | 0.28* | 2.4(1.6);9.0;n=56112 | 2.4(1.6);9.0;n=56112 | <0.01 |
| Diabetes without<br>chronic complication | 7962(14.18%) | 29884(8.39%) | 0.18 | 7962(14.18%) | 7911(14.09%) | <0.01 |
| Diabetes with chronic<br>complication | 2300(4.09%) | 5672(1.59%) | 0.15 | 2300(4.09%) | 2285(4.07%) | <0.01 |
| Hypertension | 19557(34.85%) | 61089(17.17%) | 0.41* | 19557(34.85%) | 19515(34.77%) | <0.01 |
| Heart failure | 5293(9.43%) | 19197(5.39%) | 0.15 | 5293(9.43%) | 5230(9.32%) | <0.01 |
| Atrial fibrillation | 3815(6.79%) | 14171(3.98%) | 0.12 | 3815(6.79%) | 3774(6.72%) | <0.01 |
| Renal diseases | 2529(4.50%) | 5350(1.50%) | 0.18 | 2529(4.50%) | 2512(4.47%) | <0.01 |
| Liver diseases | 211(0.37%) | 877(0.24%) | 0.02 | 211(0.37%) | 211(0.37%) | <0.01 |
| Ventricular<br>tachycardia/fibrillation | 754(1.34%) | 2141(0.60%) | 0.08 | 754(1.34%) | 754(1.34%) | <0.01 |
| Dementia and<br>Alzheimer | 622(1.10%) | 3642(1.02%) | 0.01 | 622(1.10%) | 622(1.10%) | <0.01 |
| AMI | 2781(4.95%) | 8818(2.47%) | 0.13 | 2781(4.95%) | 2772(4.94%) | <0.01 |
| COPD | 3047(5.43%) | 15278(4.29%) | 0.05 | 3047(5.43%) | 3016(5.37%) | <0.01 |
| IHD | 8958(15.96%) | 28178(7.92%) | 0.25* | 8958(15.96%) | 8849(15.77%) | 0.01 |
| PVD | 398(0.70%) | 1573(0.44%) | 0.04 | 398(0.70%) | 398(0.70%) | <0.01 |
| Stroke/TIA | 4180(7.44%) | 19768(5.55%) | 0.08 | 4180(7.44%) | 4152(7.39%) | <0.01 |
| Gastrointestinal<br>bleeding | 2852(5.08%) | 9425(2.64%) | 0.13 | 2852(5.08%) | 2820(5.02%) | <0.01 |
| Obesity | 586(1.04%) | 1227(0.34%) | 0.08 | 586(1.04%) | 586(1.04%) | <0.01 |
| ***Medications*** |  |  |  |  |  |  |
| Beta blockers | 26410(47.06%) | 163171(45.86%) | 0.02 | 26410(47.06%) | 25357(45.18%) | 0.04 |
| Calcium channel<br>blockers | 31237(55.66%) | 201941(56.76%) | 0.02 | 31237(55.66%) | 31781(56.63%) | 0.02 |
| Diuretics | 20433(36.41%) | 148940(41.86%) | 0.11 | 20433(36.41%) | 23905(42.60%) | 0.13 |

AMI: acute myocardial infarction, COPD: chronic obstructive pulmonary disease, IHD: ischemic heart disease, PVD: peripheral vascular disease, TIA: transient ischemic attack, ACEI: angiotensin-converting-enzyme inhibitors, ARB: angiotensin II receptor blockers.

**Table 2.** Significant predictors of new onset pancreatic cancer with univariate Cox analysis before and after 1:1 propensity score matching. * for p≤ 0.05, ** for p ≤ 0.01, *** for p ≤ 0.001

| Characteristics | Before Matching | P value | AfterMatching | P value |
| --- | --- | --- | --- | --- |
|  | HR[95% CI] |  | HR[95% CI] |  |
| *Demographics* |  |  |  |  |
| Male gender | 1.04[0.93, 1.16] | 0.5320 | 1.45[1.12, 1.86] | 0.0045** |
| Female gender | 1.00[reference] | - | 1.00[reference] | - |
| Baseline age, years | 1.03[1.02, 1.03] | <0.0001*** | 1.02[1.01, 1.03] | 0.0011** |
| <40 | 0.12[0.05, 0.29] | <0.0001*** | - | - |
| [40,49] | 0.30[0.21, 0.43] | <0.0001*** | 0.32[0.13, 0.78] | 0.0120* |
| [50,59] | 0.71[0.60, 0.85] | 0.0002*** | 0.86[0.60, 1.25] | 0.4290 |
| [60,69] | 1.00[reference] | - | 1.00[reference] | - |
| [70,79] | 1.34[1.19, 1.51] | <0.0001*** | 1.46[1.12, 1.90] | 0.0053** |
| >80 | 1.40[1.22, 1.62] | <0.0001*** | 1.02[1.74, 1.40] | 0.0031** |
| *Past comorbidities* |  |  |  |  |
| Charlson score | 1.14[1.10, 1.19] | <0.0001*** | 1.03[0.96, 1.11] | 0.3840 |
| CHA-DS-VASc score | 1.08[1.04, 1.13] | 0.0001*** | 0.96[0.88, 1.04] | 0.2960 |
| Diabetes without chronic complication | 1.22[1.01, 1.48] | 0.0394* | 1.21[0.85, 1.72] | 0.2830 |
| Diabetes with chronic complication | 0.90[0.55, 1.45] | 0.6520 | 0.70[0.31, 1.56] | 0.3800 |
| Hypertension | 0.90[0.77, 1.05] | 0.1850 | 0.77[0.58, 1.03] | 0.0786. |
| Heart failure | 0.81[0.59, 1.12] | 0.2020 | 0.85[0.51, 1.41] | 0.5240 |
| Atrial fibrillation | 1.09[0.80, 1.48] | 0.6030 | 1.23[0.75, 2.02] | 0.4110 |
| Renal diseases | 1.06[1.01, 1.44] | 0.0056** | 1.21[1.05, 1.86] | 0.0303* |
| Liver diseases | 1.41[0.45, 4.39] | 0.5510 | - | - |
| Ventricular tachycardia/fibrillation | 1.16[0.55, 2.43] | 0.7040 | 1.51[0.56, 4.05] | 0.4170 |
| Dementia and Alzheimer | 0.96[0.46, 2.03] | 0.9230 | - | - |
| AMI | 0.99[0.68, 1.45] | 0.9710 | 1.03[0.56, 1.88] | 0.9320 |
| COPD | 0.82[0.57, 1.18] | 0.2910 | 0.64[0.30, 1.36] | 0.2500 |
| IHD | 0.87[0.70, 1.09] | 0.2390 | 0.73[0.49, 1.09] | 0.1200 |
| PVD | 0.54[0.13, 2.15] | 0.3800 | - | - |
| Stroke/TIA | 0.66[0.48, 0.91] | 0.0113* | 0.68[0.37, 1.24] | 0.2090 |
| Gastrointestinal bleeding | 1.11[0.76, 1.61] | 0.5990 | 1.07[0.59, 1.97] | 0.8160 |
| Obesity | 0.63[0.20, 1.97] | 0.4290 | 0.42[0.06, 2.97] | 0.3830 |
| ***Medications*** |  |  |  |  |
| ARB v.s. ACEI | 0.66[0.53, 0.82] | 0.0002*** | 0.69[0.53, 0.90] | 0.0065** |
| Beta blockers | 0.89[0.80, 1.00] | 0.0548. | 0.78[0.60, 1.00] | 0.0537. |
| Calcium channel blockers | 1.02[0.91, 1.15] | 0.7220 | 1.10[0.85, 1.42] | 0.4700 |
| Diuretics | 0.96[0.85, 1.07] | 0.4410 | 1.04[0.80, 1.34] | 0.7870 |

AMI: acute myocardial infarction, COPD: chronic obstructive pulmonary disease, IHD: ischemic heart disease, PVD: peripheral vascular disease, TIA: transient ischemic attack, ACEI: angiotensin-converting-enzyme inhibitors, ARB: angiotensin II receptor blockers.

**Figure 1.**
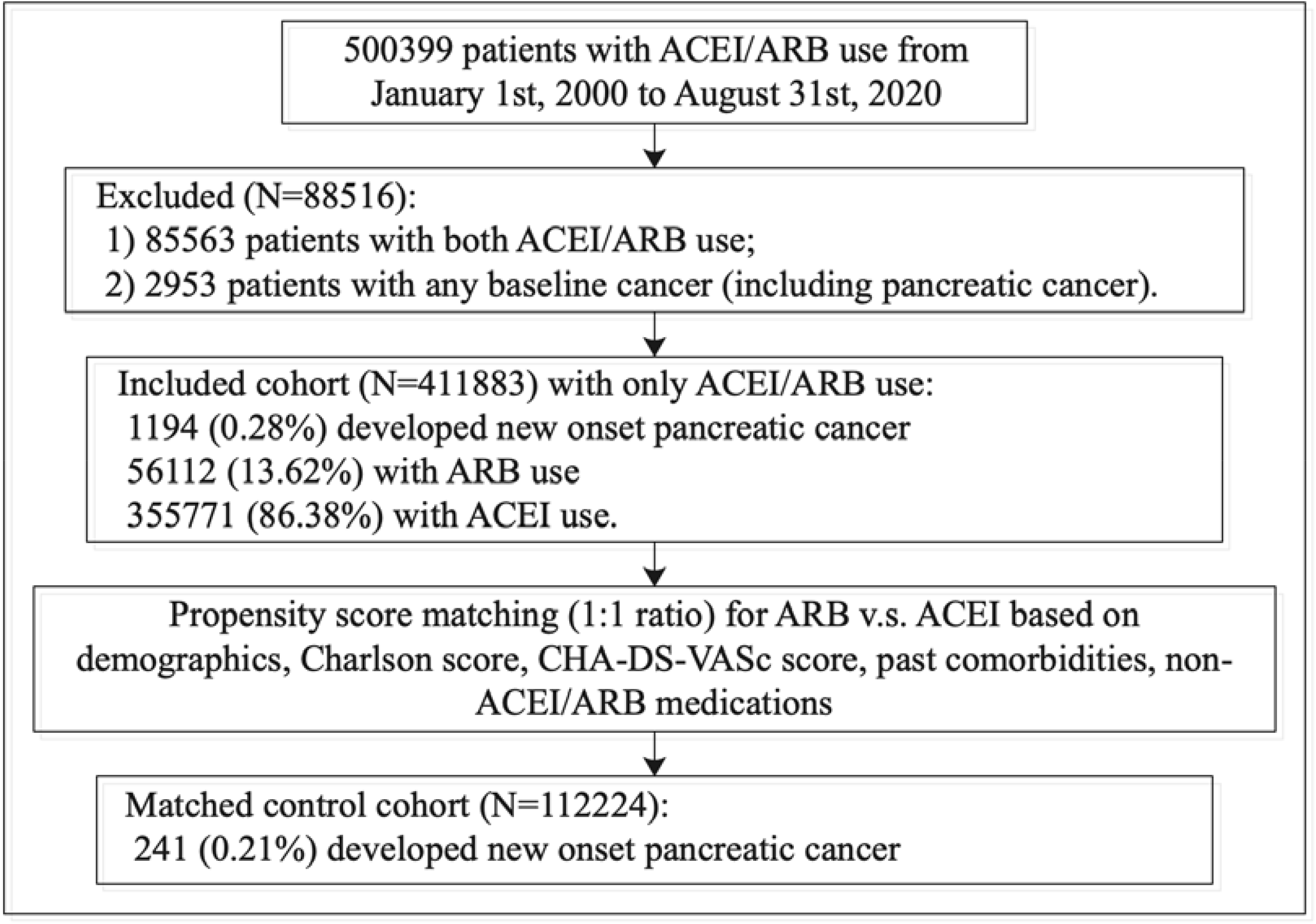
Main procedures of data processing.

### Cox regression and competing risk analysis

The number of incident cases of pancreatic cancer were higher in ACEI users compared to ARB users before (0.31% v.s. 0.15%, P<0.0001) and after propensity score matching (0.27% v.s. 0.15%, P<0.0001). Univariate Cox regression identified risk predictors in the matched cohort (**Table 3**): male gender (HR: 1.45, 95% CI: [1.12, 1.86], P=0.0045), older age at baseline as a continuous variable (HR: 1.02, 95% CI: [1.01, 1.03], P=0.0011) and age 70-80 (HR: 1.46, 95% CI: [1.12, 1.90], P=0.0053) or above 80 years old (HR: 1.02, 95% CI: [1.74, 1.40], P =0.0031), and renal disease (HR: 1.21, 95% CI: [1.05, 1.86], P=0.0303). By contrast, ARB use a lower risk of the primary outcome compared to ACEI users (HR: 0.69, 95% CI: [0.53, 0.90], P=0.0065). Cumulative incidence curves and Kaplan-Meier curves of all-cause mortality stratified by drug use of ARB v.s. ACEI before and after 1:1 propensity score matching are shown in **Figure 2** (log-rank test, P<0.05), and for new onset pancreatic cancer in **Figure 3** (log-rank test, P<0.05).

**Table 3.**
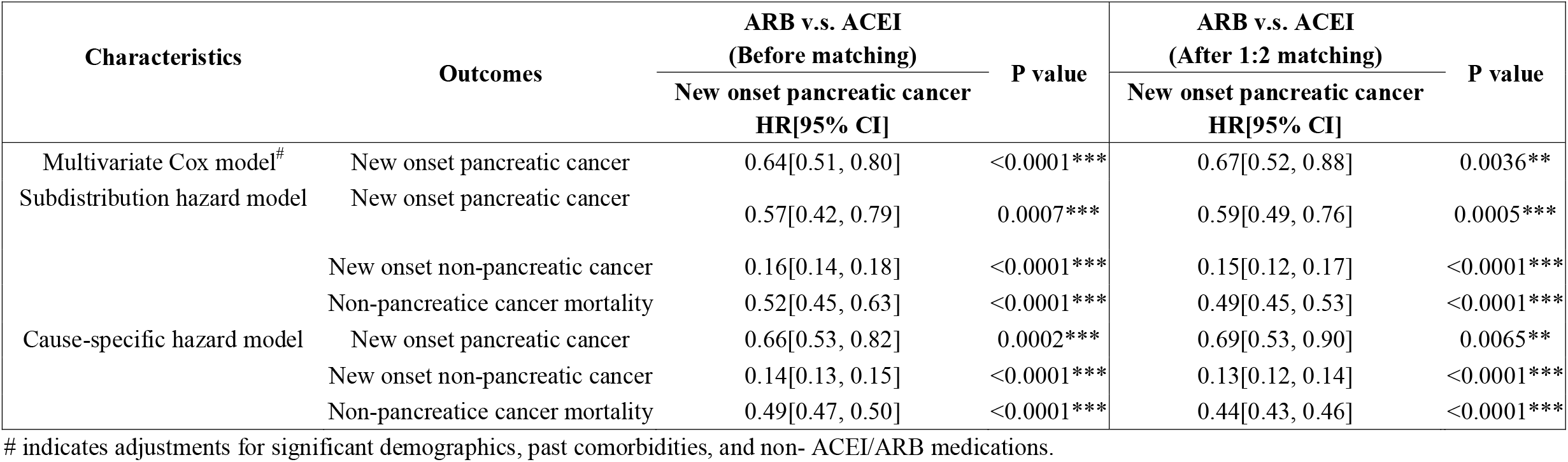
Hazard ratios (and 95% confidence intervals) from multivariate Cox models, cause-specific models and subdistribution hazard models for new onset pancreatic cancer before and after 1:1 propensity score matching. * for p≤ 0.05, ** for p ≤ 0.01, *** for p ≤ 0.001;

**Figure 2.**
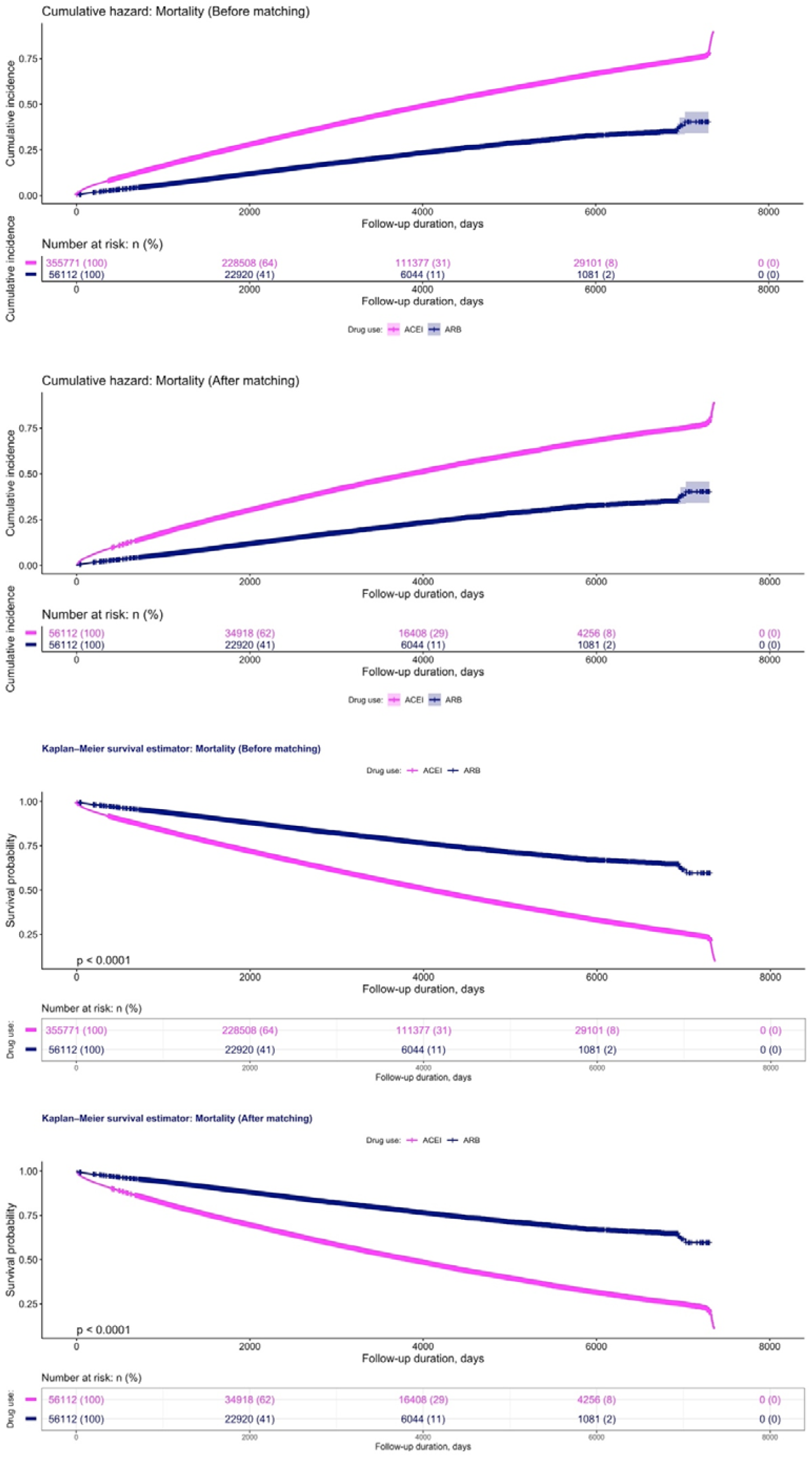
Cumulative incidence curves and Kaplan-Meier curves of all-cause mortality stratified by drug use of ARB v.s. ACEI before and after 1:1 propensity score matching.

**Figure 3.**
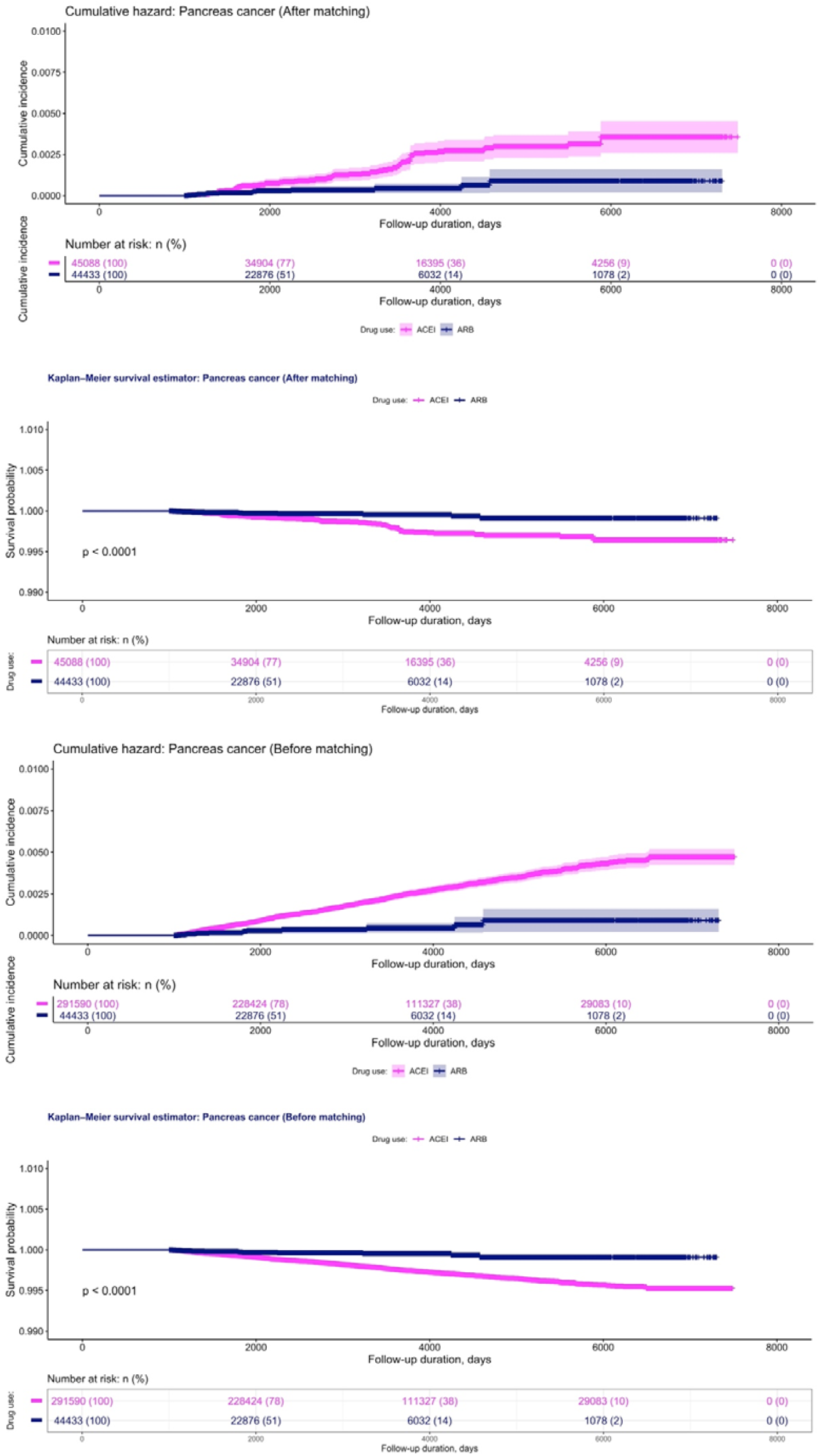
Cumulative incidence curves and Kaplan-Meier curves of new onset pancreatic cancer stratified by drug use of ARB v.s. ACEI before and after 1:1 propensity score matching.

**Figure 4.**
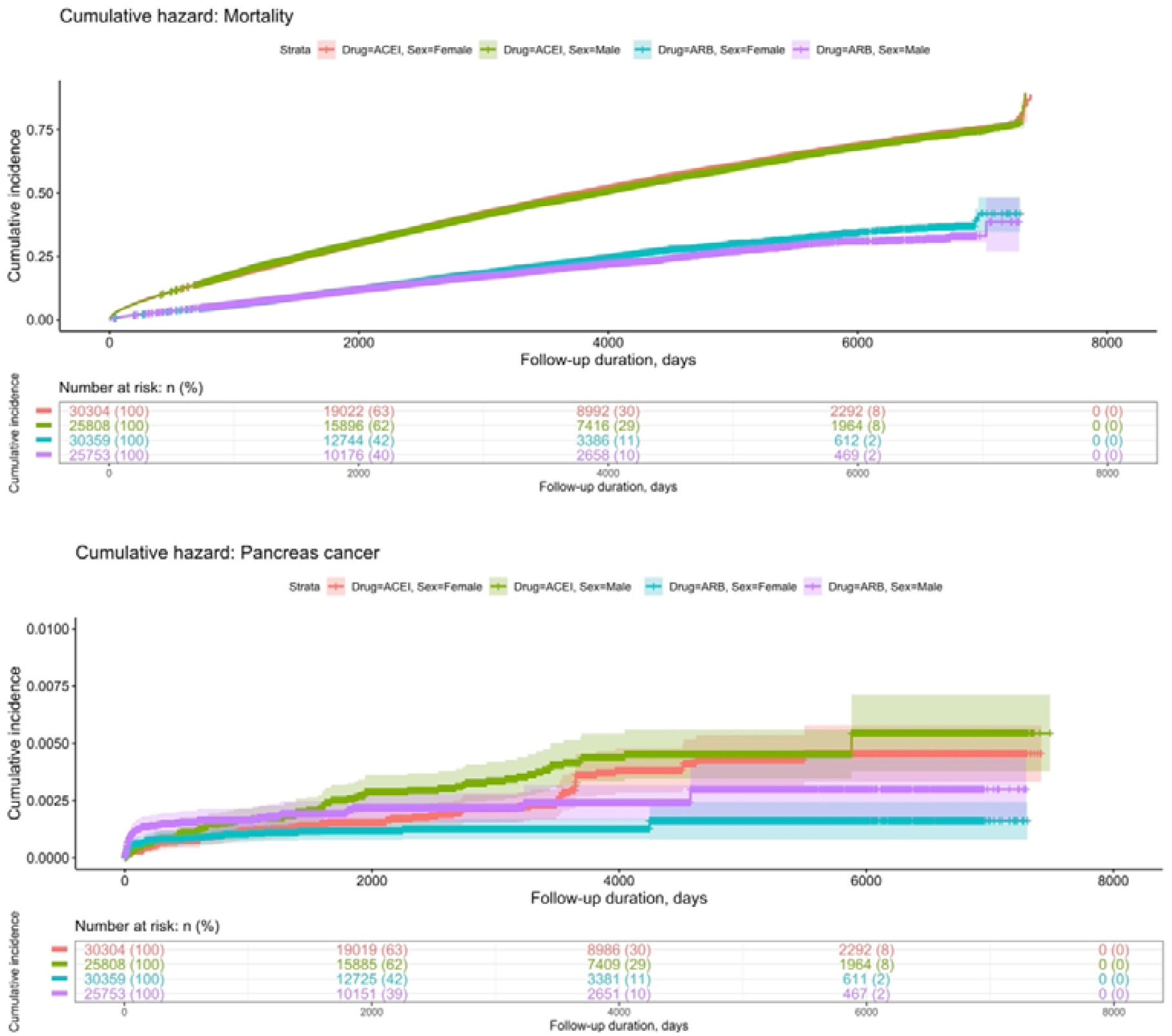
Subgroup analysis: Cumulative incidence curves of all-cause mortality and new onset pancreatic cancer stratified by the interactions between gender and drug use of ARB v.s. ACEI in the propensity score matched cohort.

The association between ARB use and lower risks of incident pancreatic cancer remained significant after multivariable adjustments (**Table 3**). Similarly, competing risk analysis using either the subdistribution hazard model (HR: 0.59, 95% CI: [0.49, 0.76], P value=0.0005) and cause-specific hazard model (HR: 0.69, 95% CI: [0.53, 0.90], P value=0.0065) confirmed this association. The cumulative incidence function is shown in **Figure 5**.

**Figure 5.**
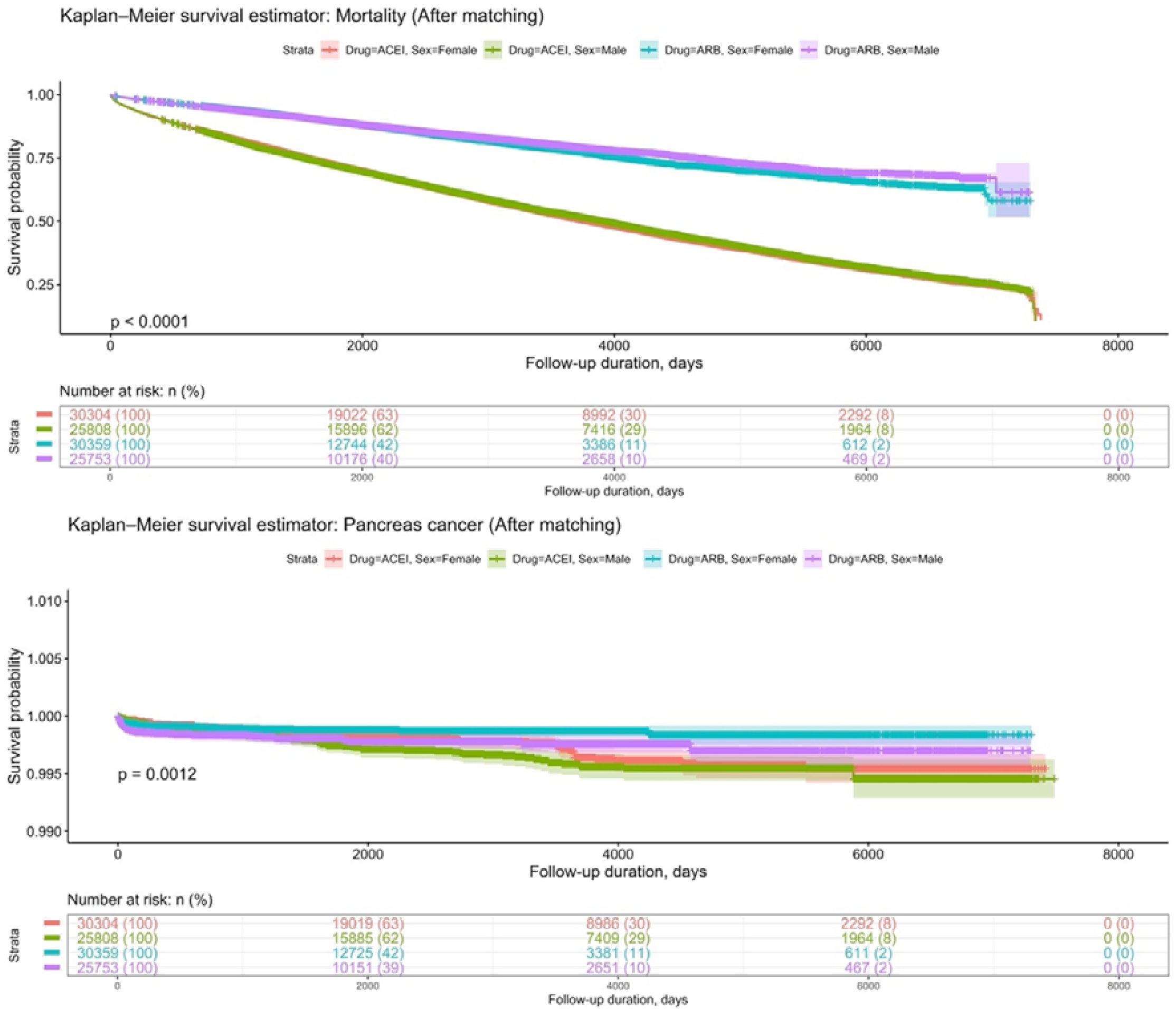
Subgroup analysis: Kaplan-Meier curves of new onset pancreatic cancer stratified by the interactions between gender and drug use of ARB v.s. ACEI in the propensity score matched cohort.

**Figure 6.**
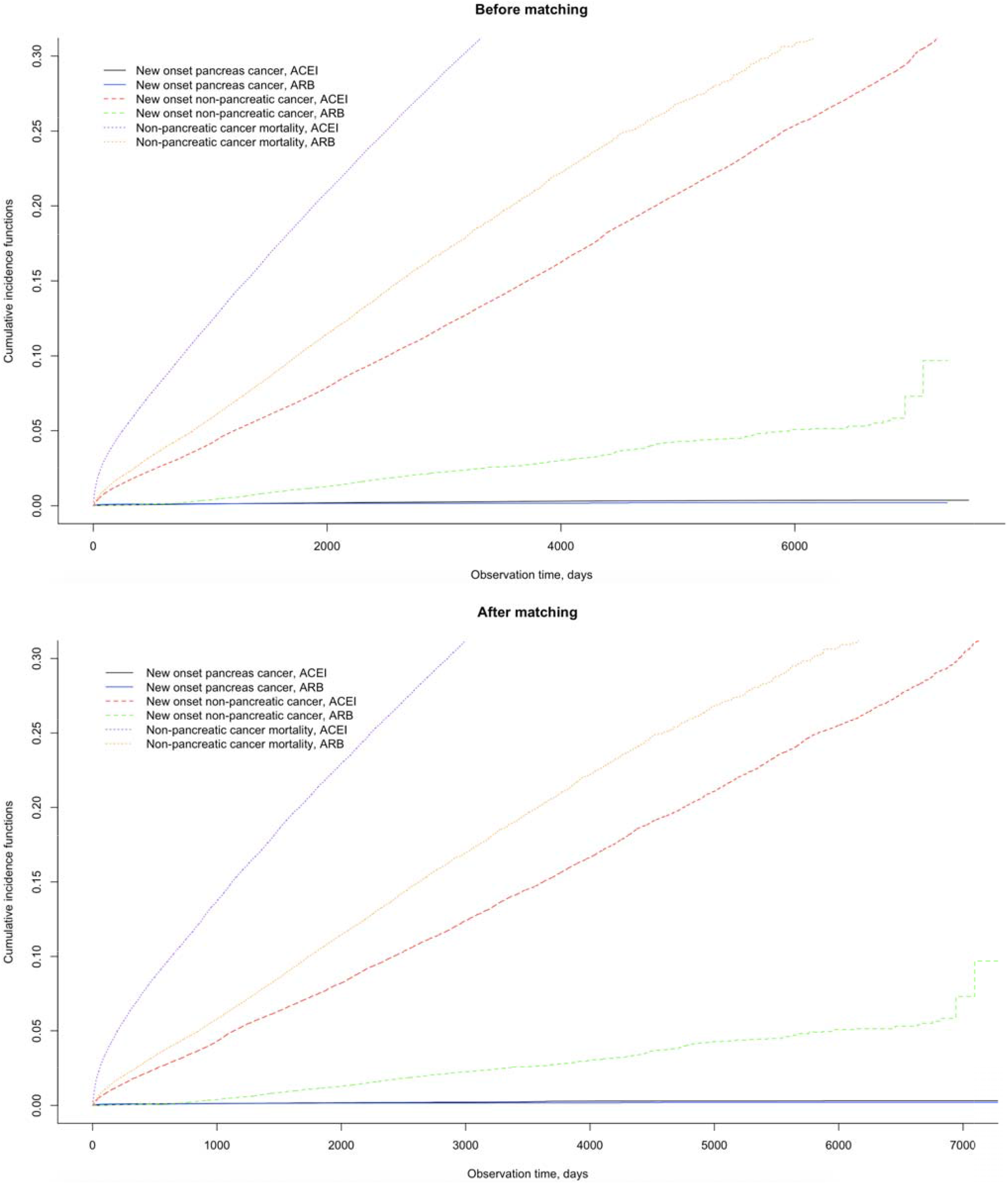
Unbiased estimates of the cumulative incidence functions in competing survival risk analysis for new onset pancreatic cancer, non-pancreatic cancers and non-pancreatic cancer related mortality stratified by ARB and ACEI monotherapy before and after 1:1 propensity score matching.

In addition, multiple propensity matching approaches were conducted to uncover the risk of incident adverse new onset pancreatic cancer and mortality risks in the matched cohorts associated with ARB v.s. ACEI exposure as presented in **Table 4**, which confirmed the same observation (HR<1, P value<0.0001).

**Table 4.** Risk of incident adverse new onset pancreatic cancer and mortality risks in the matched cohorts associated with ARB v.s. ACEI exposure using multiple propensity matching approaches. * for p≤ 0.05, ** for p ≤ 0.01, *** for p ≤ 0.001; HR: hazard ratio; CI: confidence interval; PS: propensity score, HDPS: high dimensional propensity score, IPTW: inverse probability of treatment weighting.

| Outcome | HR after PS stratification<br>[95% CI];P value | HR after HDPS matching<br>[95% CI];P value | HR after PS IPTW<br>[95% CI]; P value |
| --- | --- | --- | --- |
| New onset pancreatic cancer | 0.61[0.52-0.77];<0.0001*** | 0.65[0.55-0.75];<0.0001*** | 0.65[0.53-0.77];<0.0001*** |
| New onset non-pancreatic cancer | 0.16[0.13-0.17];<0.0001*** | 0.18[0.11-0.22];<0.0001*** | 0.17[0.15-0.19];<0.0001*** |
| Non-pancreatic cancer mortality | 0.51[0.47-0.56];<0.0001*** | 0.55[0.51-0.57];<0.0001*** | 0.54[0.51-0.56];<0.0001*** |

### Subgroup analysis: Interactions effects between gender and adverse outcomes

Cumulative incidence and Kaplan-Meier survival curves of all-cause mortality and new onset pancreatic cancer stratified by the interactions between gender and drug use of ARB v.s. ACEI in the propensity score matched cohort are presented in **Figure 5**. The results confirm the previous findings of lower pancreatic cancer risks in females for ARB exposure, whilst higher risks of pancreatic cancers were observed for ACEI in males (log-rank test, P<0.05).

## Discussion

The main finding of this study is that ARB use is associated with lower risks of incident pancreatic cancer both before and after propensity score matching, which were confirmed in competing risk analyses and inverse probability treatment weighting analysis.

RAAS inhibitors are known to modulate cell proliferation and gene expression in pancreatic cancer cells ^23^. The cytostatic effects on cancer cells of RAAS inhibitors have been attributed to their anti-oxidant actions ^24^. Other mechanisms include modulation of the ACE and angiotensin type 1 receptors in relation to angiogenesis ^25, 26^ or inhibition of growth factor signalling such as vascular epidermal growth factor/extracellular signal-regulated kinase 1/2 (ERK1/2) pathway ^27^. Moreover, ACE inhibition was shown to prevent pancreatic cancer and progression of the precursor lesion, pancreatic intraepithelial neoplasia, in a genetically engineered murine model ^28^. Additionally, it has been proposed that RAAS inhibition can affect collagen synthesis ^29^, a key element that participates in tumour growth and cancer cell proliferation ^30^. A study conducted by Chauhan *et al*. has found that losartan reduced synthesis of stromal collagen I and hyaluronan, interestingly although lisinopril exerted a similar effect, the reduction was found to be in a lesser extent compared to losartan ^31^. This may explain our finding that ARB users has a lower risk in developing pancreatic cancer than ACEI users. At a molecular level, RAAS inhibitors act via inhibiting TSP-1 which in turns reduce the downstream TGF-β signalling ^32^, and it is known to be a primary driver in promoting cancer progression by inducing epithelial–mesenchymal transition (EMT) ^33^, which is implicated in pancreatic cancer metastasis ^34^. Partly due to the pre-clinical work on the protective effects of RAAS inhibitors on carcinogenesis, clinical studies have focused on the beneficial actions of RAAS inhibitors on progression of the cancer and survival in patients with pancreatic cancer. Our findings of reduced pancreatic incidence of ARBs are in keeping with cell and animal studies that have shown that RAAS inhibition can prevent cancer development, growth and progression ^35, 36^.

### Strengths and Limitations

This study has several strengths. Firstly, this was a population-based study using the local electronic health records system, which includes patients from healthcare facilities in a single city of China. Secondly, with a study cohort of more than 400 000 patients, this is the largest study of patients prescribed ACEI and ARB with assessments of pancreatic cancer outcomes over long durations of follow-up. However, some limitations should be noted. Firstly, our study relied on administrative coding and confounders such as food intake, cigarette use, and body mass index are not routinely coded into structured data and could not be included as risk variables. Secondly, compliance to medications could not be assessed.

## Conclusions

Compared with ACEI, ARB monotherapy was linked with lower risks of new onset pancreatic cancer in unmatched and matched cohorts using Kaplan-Meier, univariable and multivariable Cox, and competing risk analyses.

## Data Availability

All data produced in the present study are available upon reasonable request to the authors

## Author Contributions

## Data availability

## References

1. Rhodes DR, Ateeq B, Cao Q, et al. AGTR1 overexpression defines a subset of breast cancer and confers sensitivity to losartan, an AGTR1 antagonist. Proc Natl Acad Sci U S A. 2009;106: 10284–10289.

2. Yang J, Yang X, Gao L, Zhang J, Yi C, Huang Y. The role of the renin-angiotensin system inhibitors in malignancy: a review. Am J Cancer Res. 2021;11: 884–897.

3. Zhou Q, Chen D-S, Xin L, et al. The renin-angiotensin system blockers and survival in digestive system malignancies: A systematic review and meta-analysis. Medicine. 2020;99: e19075–e19075.

4. Nakai Y, Isayama H, Ijichi H, et al. Inhibition of renin-angiotensin system affects prognosis of advanced pancreatic cancer receiving gemcitabine. Br J Cancer. 2010;103: 1644–1648.

5. Khoshghamat N, Jafari N, Toloue-Pouya V, et al. The therapeutic potential of renin-angiotensin system inhibitors in the treatment of pancreatic cancer. Life Sci. 2021;270: 119118.

6. Hicks BM, Filion KB, Yin H, Sakr L, Udell JA, Azoulay L. Angiotensin converting enzyme inhibitors and risk of lung cancer: population based cohort study. BMJ. 2018;363: k4209.

7. Brasky TM, Flores KF, Larson JC, et al. Associations of Angiotensin-Converting Enzyme Inhibitor or Angiotensin Receptor Blocker Use with Colorectal Cancer Risk in the Women’s Health Initiative. Cancer Epidemiol Biomarkers Prev. 2021.

8. Chen X, Yi CH, Ya KG. Renin-angiotensin system inhibitor use and colorectal cancer risk and mortality: A dose-response meta analysis. J Renin Angiotensin Aldosterone Syst. 2020;21: 1470320319895646.

9. Kirkegard J, Mortensen FV, Cronin-Fenton D. Antihypertensive drugs and pancreatic cancer risk in patients with chronic pancreatitis: a Danish nationwide population-based cohort study. Br J Cancer. 2019;121: 622–624.

10. Wang Z, White DL, Hoogeveen R, et al. Anti-Hypertensive Medication Use, Soluble Receptor for Glycation End Products and Risk of Pancreatic Cancer in the Women’s Health Initiative Study. J Clin Med. 2018;7.

11. Cerullo M, Gani F, Chen SY, Canner JK, Pawlik TM. Impact of Angiotensin Receptor Blocker Use on Overall Survival Among Patients Undergoing Resection for Pancreatic Cancer. World J Surg. 2017;41: 2361–2370.

12. Mandilaras V, Bouganim N, Yin H, Asselah J, Azoulay L. The use of drugs acting on the renin-angiotensin system and the incidence of pancreatic cancer. Br J Cancer. 2017;116:103–108.

13. Ju C, Lai RWC, Li KHC, et al. Comparative cardiovascular risk in users versus non-users of xanthine oxidase inhibitors and febuxostat versus allopurinol users. Rheumatology (Oxford). 2020;59: 2340–2349.

14. Zhou J, Wang X, Lee S, et al. Proton pump inhibitor or famotidine use and severe COVID-19 disease: a propensity score-matched territory-wide study. Gut. 2020.

15. Tse G, Zhou J, Lee S, et al. Relationship between angiotensin-converting enzyme inhibitors or angiotensin receptor blockers and COVID-19 incidence or severe disease. J Hypertens. 2021;39: 1717–1724.

16. Zhou J, Tse G, Lee S, et al. Interaction effects between angiotensin-converting enzyme inhibitors or angiotensin receptor blockers and steroid or anti-viral therapies in COVID-19: a population-based study. J Med Virol. 2021.

17. Zhou J, Lee S, Wang X, et al. Development of a multivariable prediction model for severe COVID-19 disease: a population-based study from Hong Kong. NPJ Digit Med. 2021;4: 66.

18. Lee S, Zhou J, Guo CL, et al. Predictive scores for identifying patients with type 2 diabetes mellitus at risk of acute myocardial infarction and sudden cardiac death. Endocrinology, Diabetes & Metabolism. 2021;n/a: e00240.

19. Lee S, Liu T, Zhou J, Zhang Q, Wong WT, Tse G. Predictions of diabetes complications and mortality using hba1c variability: a 10-year observational cohort study. Acta Diabetol. 2021;58: 171–180.

20. Austin PC. An Introduction to Propensity Score Methods for Reducing the Effects of Confounding in Observational Studies. Multivariate Behav Res. 2011;46: 399–424.

21. Schneeweiss S, Rassen JA, Glynn RJ, Avorn J, Mogun H, Brookhart MA. High-dimensional propensity score adjustment in studies of treatment effects using health care claims data. Epidemiology. 2009;20: 512–522.

22. Austin PC, Stuart EA. Moving towards best practice when using inverse probability of treatment weighting (IPTW) using the propensity score to estimate causal treatment effects in observational studies. Stat Med. 2015;34: 3661–3679.

23. Reddy MK, Baskaran K, Molteni A. Inhibitors of angiotensin-converting enzyme modulate mitosis and gene expression in pancreatic cancer cells. Proc Soc Exp Biol Med. 1995;210: 221–226.

24. Molteni A, Ward WF, Ts’ao CH, et al. Cytostatic properties of some angiotensin I converting enzyme inhibitors and of angiotensin II type I receptor antagonists. Curr Pharm Des. 2003;9: 751–761.

25. Arafat HA, Gong Q, Chipitsyna G, Rizvi A, Saa CT, Yeo CJ. Antihypertensives as novel antineoplastics: angiotensin-I-converting enzyme inhibitors and angiotensin II type 1 receptor blockers in pancreatic ductal adenocarcinoma. J Am Coll Surg. 2007;204: 996-1005; discussion 1005-1006.

26. Noguchi R, Yoshiji H, Ikenaka Y, et al. Synergistic inhibitory effect of gemcitabine and angiotensin type-1 receptor blocker, losartan, on murine pancreatic tumor growth via anti-angiogenic activities. Oncol Rep. 2009;22: 355–360.

27. Anandanadesan R, Gong Q, Chipitsyna G, Witkiewicz A, Yeo CJ, Arafat HA. Angiotensin II induces vascular endothelial growth factor in pancreatic cancer cells through an angiotensin II type 1 receptor and ERK1/2 signaling. J Gastrointest Surg. 2008;12: 57–66.

28. Fendrich V, Chen NM, Neef M, et al. The angiotensin-I-converting enzyme inhibitor enalapril and aspirin delay progression of pancreatic intraepithelial neoplasia and cancer formation in a genetically engineered mouse model of pancreatic cancer. Gut. 2010;59: 630–637.

29. Pallasch FB, Schumacher U. Angiotensin Inhibition, TGF-beta and EMT in Cancer. Cancers (Basel). 2020;12.

30. Xu S, Xu H, Wang W, et al. The role of collagen in cancer: from bench to bedside. J Transl Med. 2019;17: 309.

31. Chauhan VP, Martin JD, Liu H, et al. Angiotensin inhibition enhances drug delivery and potentiates chemotherapy by decompressing tumour blood vessels. Nat Commun. 2013;4: 2516.

32. Naito T, Masaki T, Nikolic-Paterson DJ, Tanji C, Yorioka N, Kohno N. Angiotensin II induces thrombospondin-1 production in human mesangial cells via p38 MAPK and JNK: a mechanism for activation of latent TGF-beta1. Am J Physiol Renal Physiol. 2004;286: F278–287.

33. Katsuno Y, Lamouille S, Derynck R. TGF-beta signaling and epithelial-mesenchymal transition in cancer progression. Curr Opin Oncol. 2013;25: 76–84.

34. Rhim AD, Mirek ET, Aiello NM, et al. EMT and dissemination precede pancreatic tumor formation. Cell. 2012;148: 349–361.

35. Liu H, Naxerova K, Pinter M, et al. Use of Angiotensin System Inhibitors Is Associated with Immune Activation and Longer Survival in Nonmetastatic Pancreatic Ductal Adenocarcinoma. Clinical Cancer Research. 2017;23: 5959–5969.

36. Pinter M, Jain RK. Targeting the renin-angiotensin system to improve cancer treatment: Implications for immunotherapy. Science Translational Medicine. 2017;9: eaan5616.

