## Supplementary Appendix for "Incidence of pancreatic cancer in angiotensin-converting enzyme inhibitors (ACEIs) versus angiotensin receptor blockers (ARBs): a population-based cohort study"

**Supplementary Table 1. Codes for past comorbidities**

Diabetes without chronic complication 250	250.01	250.02	250.03	250.1	250.11	250.12	250.13	250.2	250.21	250.22	250.23	250.3	250.31	250.32	250.33	250.7	250.71	250.72	250.73

Diabetes with chronic complication	250.4	250.41	250.42	250.43	250.5	250.51	250.52	250.53	250.6	250.61	250.62	250.63

Renal diseases	582	582	582.1	582.2	582.4	582.8	582.81	582.89	582.9	583	583	583.1	583.2	583.4	583.6	583.7	585	585.1	585.2	585.3	585.4	585.5	585.6	585.9	586	588	588	588.1	588.8	588.81	588.89	588.9

Systemic embolism	444	444.01	444.09	444.1	444.2	444.21	444.22	444.8	444.81	444.89	444.9	445	445.01	445.02	445.8	445.81	445.89

Hypertension	401	401.1	401.9	402	402.01	402.1	402.11	402.9	402.91	403	403.01	403.1	403.11	403.9	403.91	404	404.01	404.02	404.03	404.1	404.11	404.12	404.13	404.9	404.91	404.92	404.93	405	405.01	405.09	405.1	405.11	405.19	405.9	405.91	405.99	437.2

Heart failure	428	428	428.1	428.2	428.2	428.21	428.22	428.23	428.3	428.3	428.31	428.32	428.33	428.4	428.4	428.41	428.42	428.43	428.9	398.91	402.01	402.11	402.91	404.01	404.03	404.11	404.13	404.91	404.93

Atrial fibrillation	427.31	429.4

Ventricular arrhythmias/sudden cardiac death 410	410.01	410.02	410.1	410.11	410.12	410.2	410.21	410.22	410.3	410.31	410.32	410.4	410.41	410.42	410.5	410.51	410.52	410.6	410.61	410.62	410.7	410.71	410.72	410.8	410.81	410.82	410.9	410.91	410.92	427.01	427.1	427.4	427.4	427.41	427.42	427.5	427.5	427.69	798	798.1	798.2

Liver diseases	456	456.1	456.2	572.2	572.3	572.4	572.8	571.4	571.5	571.6

Dementia	331.82	290	290.1	290.11	290.12	290.13	290.2	290.21	290.3	290.4	290.41	290.42	290.43	290.8	290.9	294.2	294.1	294.11	294.21	332	46.1	333.4	340	42331	331.19	294.29

Chronic obstructive pulmonary disease	490	491	492	493	494	495	496	491.1	491.2	491.21	491.22	491.8	491.9	492.8	493.01	493.02	493.1	493.11	493.12	493.2	493.21	493.22	493.8	493.81	493.82	493.9	493.91	493.92	494.1	495.1	495.2	495.3	495.4	495.5	495.6	495.7	495.8	495.9

Peripheral vascular disease	250.7	443.9	443	443.1	443.2	443.21	443.22	443.23	443.24	443.29	443.8	443.81	443.82	443.89	441	443.9	785.4	V43.4

Stroke/transient ischemic attack	Stroke/TIA	435	435.1	435.2	435.3	435.8	435.9	433.81	433.91	434	436	437	437.1	433.31	433.01	434.01	434.1	434.11	434.9	434.91	437.2	437.3	437.4	437.5	437.6	437.7	437.8	437.9	430	431	432	432.1	432.9

Gastrointestinal bleeding	531	531.2	531.4	531.6	532	532.2	532.4	532.6	533	533.2	533.4	533.6	534	534.2	534.4	534.6	535.01	535.11	535.21	535.31	535.41	535.51	535.61	535.71	562.02	562.03	562.12	562.13	569.3	569.85	569.86	578	578.1	578.9

Ischemic heart disease	410.01	410.02	410.1	410.11	410.12	410.2	410.21	410.22	410.3	410.31	410.32	410.4	410.41	410.42	410.5	410.51	410.52	410.6	410.61	410.62	410.7	410.71	410.72	410.8	410.81	410.82	410.9	410.91	410.92	411	411.1	411.8	411.81	411.89	413	413.1	413.9	414	414.01	414.02	414.03	414.04	414.05	414.06	414.07	414.1	414.11	414.12	414.19	414.2	414.3	414.4	414.8	414.9	410	412

Cancer 140-239

Obesity	278.01	278	278

**Supplementary Table 2. Baseline characteristics of patients with/without new onset pancreatic cancer after 1:1 propensity score matching.**

* for SMD$\geq$0.02; # indicates the difference between patients with/without new onset pancreatic cancer

| **Characteristics** | **Before matching** |  | **SMD** | **After matching** |  | **SMD** |
| --- | --- | --- | --- | --- | --- | --- |
|  | **New onset pancreas cancer (N=1194) Mean(SD);Max;N or Count(%)** | **No new onset pancreas cancer (N=410689) Mean(SD);Max;N or Count(%)** |  | **New onset pancreas cancer (N=241) Mean(SD);Max;N or Count(%)** | **No new onset pancreas cancer (N=111983) Mean(SD);Max;N or Count(%)** |  |
| ***Demographics*** |  |  |  |  |  |  |
| Male gender | 640(53.60%) | 217820(53.03%) | 0.01 | 132(54.77%) | 51429(45.92%) | 0.18 |
| Female gender | 554(46.39%) | 192869(46.96%) | 0.01 | 109(45.22%) | 60554(54.07%) | 0.18 |
| Baseline age, years | 70.8(10.7);100.6;n=1194 | 69.2(13.8);116.3;n=410689 | 0.13 | 70.8(10.9);95.0;n=241 | 69.5(13.6);110.2;n=111983 | 0.1 |
| <40 | 5(0.41%) | 11014(2.68%) | 0.18 | 0(0.00%) | 2510(2.24%) | 0.21* |
| [40,49] | 28(2.34%) | 24656(6.00%) | 0.18 | 5(2.07%) | 5977(5.33%) | 0.17 |
| [50,59] | 139(11.64%) | 54854(13.35%) | 0.05 | 33(13.69%) | 15810(14.11%) | 0.01 |
| [60,69] | 270(22.61%) | 81599(19.86%) | 0.07 | 55(22.82%) | 23982(21.41%) | 0.03 |
| [70,80] | 422(35.34%) | 119952(29.20%) | 0.13 | 83(34.43%) | 29386(26.24%) | 0.18 |
| >80 | 248(20.77%) | 95915(23.35%) | 0.06 | 50(20.74%) | 28166(25.15%) | 0.1 |
| ***Past comorbidities*** |  |  |  |  |  |  |
| Charlson score | 2.9(1.3);8.0;n=1194 | 2.9(1.6);15.0;n=410689 | 0 | 3.0(1.3);7.0;n=241 | 3.1(1.8);14.0;n=111983 | 0.09 |
| CHA-DS-VASc score | 1.9(1.4);7.0;n=1194 | 2.0(1.5);9.0;n=410689 | 0.05 | 2.1(1.5);7.0;n=241 | 2.4(1.6);9.0;n=111983 | 0.17 |
| Diabetes without chronic complication | 117(9.79%) | 37729(9.18%) | 0.02 | 37(15.35%) | 15836(14.14%) | 0.03 |
| Diabetes with chronic complication | 17(1.42%) | 7955(1.93%) | 0.04 | 6(2.48%) | 4579(4.08%) | 0.09 |
| Hypertension | 180(15.07%) | 80466(19.59%) | 0.12 | 65(26.97%) | 39007(34.83%) | 0.17 |
| Heart failure | 40(3.35%) | 24450(5.95%) | 0.12 | 16(6.63%) | 10507(9.38%) | 0.1 |
| Atrial fibrillation | 41(3.43%) | 17945(4.36%) | 0.05 | 17(7.05%) | 7572(6.76%) | 0.01 |
| Renal diseases | 1(0.08%) | 7878(1.91%) | 0.19 | 2(0.82%) | 5039(4.49%) | 0.23* |
| Liver diseases | 3(0.25%) | 1085(0.26%) | 0 | 0(0.00%) | 422(0.37%) | 0.09 |
| Ventricular tachycardia/fibrillation | 7(0.58%) | 2888(0.70%) | 0.01 | 4(1.65%) | 1504(1.34%) | 0.03 |
| Dementia and Alzheimer | 7(0.58%) | 4257(1.03%) | 0.05 | 0(0.00%) | 1244(1.11%) | 0.15 |
| AMI | 27(2.26%) | 11572(2.81%) | 0.04 | 11(4.56%) | 5542(4.94%) | 0.02 |
| COPD | 30(2.51%) | 18295(4.45%) | 0.11 | 7(2.90%) | 6056(5.40%) | 0.13 |
| IHD | 83(6.95%) | 37053(9.02%) | 0.08 | 28(11.61%) | 17779(15.87%) | 0.12 |
| PVD | 2(0.16%) | 1969(0.47%) | 0.05 | 0(0.00%) | 796(0.71%) | 0.12 |
| Stroke/TIA | 38(3.18%) | 23910(5.82%) | 0.13 | 11(4.56%) | 8321(7.43%) | 0.12 |
| Gastrointestinal bleeding | 28(2.34%) | 12249(2.98%) | 0.04 | 11(4.56%) | 5661(5.05%) | 0.02 |
| Obesity | 3(0.25%) | 1810(0.44%) | 0.03 | 1(0.41%) | 1171(1.04%) | 0.07 |
| ***Medications*** |  |  |  |  |  |  |
| ARB v.s. ACEI | 88(7.37%) | 56024(13.64%) | 0.21* | 88(36.51%) | 56024(50.02%) | 0.28* |
| Beta blockers | 516(43.21%) | 189065(46.03%) | 0.06 | 96(39.83%) | 51671(46.14%) | 0.13 |
| Calcium channel blockers | 682(57.11%) | 232496(56.61%) | 0.01 | 141(58.50%) | 62877(56.14%) | 0.05 |
| Diuretics | 480(40.20%) | 168893(41.12%) | 0.02 | 98(40.66%) | 44240(39.50%) | 0.02 |

AMI: acute myocardial infarction, COPD: chronic obstructive pulmonary disease, IHD: ischemic heart disease, PVD: peripheral vascular disease, TIA: transient ischemic attack, ACEI: angiotensin-converting-enzyme inhibitors, ARB: angiotensin II receptor blockers.
